# Hepatectomy combined with targeted and immunotherapy for CNLC stage IIIb hepatocellular carcinoma: a single-arm clinical trials protocol

**DOI:** 10.1101/2022.07.29.22278175

**Authors:** Jun-Tao Huang, Jian-Hong Zhong, Jie Zhang, Wen-Feng Gong, Liang Ma, Le-Qun Li, Bang-De Xiang

## Abstract

**Introduction:** Current clinical guidelines recommend systematic antitumor therapy as the primary treatment option for patients with stage IIIb hepatocellular carcinoma (HCC) based on the China liver cancer staging (CNLC) criteria. Several different targeted therapeutics have been applied in combination with immunotherapeutic regimens to date in patients with advanced HCC. The present study was developed to evaluate the relative safety and efficacy of hepatectomy in combination with targeted apatinib treatment and immunotherapeutic camrelizumab treatment CNLC-IIIb stage HCC patients with the goal of providing evidence regarding the potential value of this therapeutic regimen in individuals diagnosed with advanced HCC.

**Methods and analysis:** This is a single-arm multicenter clinical trial in which patients undergo hepatectomy in combination with targeted treatment (apatinib) and immunotherapy (camrelizumab). Patients will undergo follow-up every 2-3 months following treatment initiation to record any evidence of disease progression and adverse event incidence for a minimum of 24 months following the discontinuation of treatment until reaching study endpoint events or trial termination. The primary endpoint for this study is patient mortality.

**Ethics and dissemination:** This study protocol was approved by the Ethics Committee of the Guangxi Medical University Cancer Hospital for Human Study (reference number KS2022[124]). The results of this study will be submitted for publication in a peer-reviewed journal.

**Trial registration number:** NCT05062837.

**Strengths and limitations of this study:**

1. This study will be the first to assess the relative safety and efficacy of hepatectomy combined with targeted and immunotherapeutic treatment in CNLC-IIIb HCC patients.
2. As a multicenter study, the results of this analysis will be representative, generalizable, and reliable.
3. As this study will entail a prolonged follow-up period, it is critical that participants be thoroughly informed prior to enrollment, with individuals exhibiting high compliance being chosen for study inclusion.

## INTRODUCTION

Hepatocellular carcinoma (HCC) is the seventh most common form of cancer, and causes roughly 600,000 deaths annually.^[1]^ As early-stage HCC generally fails to exhibit any specific symptoms, patients are primarily diagnosed with this disease when it is already in a more advanced stage in China.^[2]^ Between 2003 and 2013, our group identified 6,241 patients with HCC in the Guangxi Medical University Cancer Hospital, with 54% of these patients exhibiting disease consistent with stage C Barcelona Clinic Liver Cancer Staging (BCLC).^[2]^ Similarly, studies conducted in Hong Kong (n = 3,856) and Italy (n = 5,183) have respectively reported stage C HCC patient proportions of 40% and 45%. ^[3, 4]^ The median survival of advanced BCLC stage C patients is just 4-6 months, even with optimal supportive treatment.^[5, 6]^ Advanced HCC cases can be further stratified into those exhibiting macrovascular invasion (CNLC IIIa) and those exhibiting extrahepatic metastases (CNLC IIIb).^[7]^ Molecular targeted drug-based treatment is currently recommended in many countries for patients with CNLC-IIIb HCC.^[7-13]^ However, the single-agent efficacy of these targeted therapeutics is generally reported to be relatively limited when used to treat advanced HCC, and patients generally exhibit unsatisfactory improvements in long-term prognostic outcomes.^[14-19]^

Immune checkpoint inhibitor (ICI) use in patients with advanced-stage HCC has been a focus of growing interest in recent years owing to evidence of their efficacy in several solid tumor types.^[20]^ Patients that undergo ICI treatment in combination with targeted therapy have been reported to exhibit significantly improved overall survival (OS) and progression-free survival (PFS) relative to patients that underwent monotherapy treatment.^[21-24]^ The combined treatment of advanced HCC patients with levatinib and ICIs has also been shown to be associated with markedly improved OS relative to monotherapy.^[25, 26]^ As such, combinations of targeted therapeutics and ICIs represent the most promising treatment options for advanced-stage HCC. Accordingly, the CFDA has approved the use of targeted drug apatinib and the PD-1 inhibitor camrelizumab as second-line treatment for HCC,^[27]^ and the effectiveness and safety of this combination therapeutic regimen when used as a first-line treatment for advanced HCC have also been confirmed.^[28]^

Local progression of intrahepatic tumors is the leading cause of death for patients with advanced HCC, which is far more lethal than extrahepatic metastases. While hepatectomy is not recommended for CNLC-IIIb HCC patients under domestic or international guidelines, ^[7-13]^ there is retrospective evidence suggesting that patients who undergo hepatectomy, including palliative resection, exhibit significantly improved treatment outcomes relative to those who do not undergo surgical treatment. Surgically treated patients reportedly exhibit a median OS of up to 32 months,^[29-32]^ with this duration being significantly longer than that achieved by patients who do not undergo surgical treatment and are instead treated with combinations of targeted drugs and ICIs.^[22, 28]^ Previous retrospective evidence further suggests that certain HCC patients exhibiting extrahepatic metastases may still exhibit long-term survival benefits following hepatectomy.^[33, 34]^

Given the above evidence, we hypothesize that surgical resection of intrahepatic tumors can benefit patients by reducing the tumor load and proinflammatory cytokine release. Afterthat, the combination of VEGF inhibitors and ICI immunotherapy can promote the shrinkage or regression of extrahepatic metastases and ultimately prolong the survival time of CNLC-IIIb HCC patients.^[35-38]^ As such, the present study protocol was formulated based on a combined regimen of apatinib plus camrelizumab as data from several studies have indicated that this combination is associated with the longest median survival time for treated patients.^[22, 24, 28]^ This study will prospectively enroll stage CNLC-IIIb HCC patients with extrahepatic metastases and which intrahepatic lesions are considered suitable for radical resection. Recruitment and analyses will be performed across multiple clinical centers in China with the goal of assessing the OS for these patients following treatment via hepatectomy in combination with apatinib plus camrelizumab and the PFS, objective response rate (ORR), disease control rate (DCR), duration of response (DOR), and time to progression (TTP) of extrahepatic lesions

## METHODS AND ANALYSIS

### Patient eligibility criteria

1. Age: 18-75 years.
2. CNLC stage IIIb HCC with extrahepatic metastases including lymph node, bone, and lung metastases, but excluding brain metastases, as diagnosed via clinical imaging based on the Liver Imaging Reporting and Data System.
3. Patients are considered candidates for the radical surgical resection of local liver tumors.
4. Liver function: Grade A Child-Pugh score.
5. Indo-Cyanine Green retention value: 15 min (ICG R15) <10%.
6. Eastern Cooperative Oncology Group-Performance status (ECOG-PS): 0 or 1.
7. Expected survival duration: ≥ 6 months^[39]^.
8. Hematological indexes that do not exceed the following thresholds: hemoglobin ≥ 90 g/L; absolute neutrophil count ≥ 1.5 × 10^9^/L; platelets ≥ 80 × 10^9^/L; total bilirubin ≤ 1.5x the upper limit of normal (ULN); alanine transaminase (ALT) < 3x ULN; aspartate aminotransferase (AST) < 3x ULN or less; alkaline phosphatase (AKP) ≤ 2.5x ULN; serum albumin ≥ 28 g/L; serum creatinine ≤ 1.5 x ULN.
9. Patients do not wish to go through systemic treatments such as radiotherapy or transcatheter arterial chemoembolization (TACE).
10. All women of childbearing age will be required to utilize contraception (e.g., condoms, contraceptive pills, or intrauterine devices) for the duration of the trial and for 3 months following trial completion and must exhibit negative serum or urinary urine human chorionic gonadotropin (HCG) test result within 72 hours prior to study enrollment. All male study participants with female partners of childbearing age will be required to utilize effective contraception throughout the study duration and for 3 months following study completion.

### Patient exclusion criteria

Patients will not be eligible for inclusion if they meet any of the following criteria:

1. A history of prior or concurrent malignancies, with the exception of cured cases of cutaneous basal cell carcinoma or carcinoma *in situ* of the cervix.
2. A history of using other chemotherapeutic or immunosuppressive drugs to treat HCC, including but not limited to toripalimab, atezolizumab, sintilimab, pembrolizumab, tislelizumab, nivolumab, adriamycin, and S-1.
3. Previous radiotherapy, TACE, or systemic treatment within the past 6 months.
4. Evidence of congenital or acquired immunodeficiency disorders, including HIV positivity.
5. A known severe allergy to PD-1 monoclonal antibody treatment.
6. A temperature ≥ 38.5□ or a white blood cell count >15 × 10^9^/L of undetermined etiology within 7 days prior to study enrollment.
7. Patients diagnosed with combined bleeding disorders within 3 months of study enrollment including but not limited to moderate/severe gastro-esophageal varices, gastrointestinal bleeding, bleeding gastric ulcers, hemoptysis (> 2.5 mL/day). In cases where patients exhibit positive fecal occult blood test results, repeat testing and gastroenteroscopy should be conducted as appropriate.
8. Individuals with a history of arterial or venous thrombotic events within 6 months prior to study enrollment, including deep vein thrombosis, pulmonary infarction, or cerebrovascular accidents including cerebral ischemia, cerebral infarction, and transient ischemic attack.
9. Patients with a history of active psychotropic drug or alcohol abuse or with diagnosed mental health disorders.
10. Females who are currently lactating.
11. Individuals with active or previously diagnosed autoimmune diseases including autoimmune hepatitis, interstitial pneumonia, uveitis, enterocolitis, hypophysitis, vasculitis, nephritis, and hyperthyroidism.
12. Reported utilization of immunosuppressive drugs or hormonal therapies within 2 weeks prior to study enrollment.
13. Most recent molecular targeted therapy use (including but not limited to: sorafenib, erlotinib, lenvatinib, donafenib, and regorafenib) for less than 5 drug half-lives or failure to recover from prior therapy-related adverse event (AE) to grade 1 of the common terminology criteria for adverse events (CTCAE).
14. Individuals exhibiting combined hepatic encephalopathy or brain metastases.
15. Patients affected by hypertension not effectively controlled with medication (systolic blood pressure ≥ 140 mmHg or diastolic blood pressure ≥ 90 mmHg).
16. Patients exhibiting uncontrolled heart disease or symptoms such as myocardial infarctions within the last 12 months, unstable angina, class II or higher cardiac function, or supraventricular or ventricular arrhythmias necessitating treatment or intervention.
17. Patients with abnormal coagulatory function (INR >2.0, PT >16s), bleeding tendencies, or a requirement for thrombolytic or anticoagulation therapy, but prophylactic use of low-dose aspirin or low-molecular-weight heparin is permitted.
18. Patients with hereditary or acquired blood disorders (e.g., hemophilia, thrombocytopenia, or coagulation disorders).
19. Individuals with urinary protein levels ≥ ++ during routine urine and 24-hour urine protein levels ≥ 1.0 g.

### Study protocol

This is a multicenter single-arm clinical trial. Investigators will introduce the study to patients deemed eligible to participate based on the above criteria, after which informed consent forms will be proffered. Those patients that provide written informed consent will be enrolled in the treatment group, and will be assigned an independent study number for data collection and follow-up. Hepatectomy will be scheduled within one week of study enrollment, with apatinib and camrelizumab therapy being initiated 2-4 weeks later.

### Recruitment

In total, this study will enroll 62 patients from Guangxi Medical University Cancer Hospital and other hospitals. Recruitment began in November 2021 and is expected to end in January 2023.

### Trial intervention

Patients who, following clinical evaluation, are considered to be able to tolerate hepatectomy will undergo radical liver tumor resection while intubated under general anesthesia. For patients with lymph node and abdominal metastases, lymph nodes should be removed intraoperatively if patients exhibit other target lesions that can be used to observe the efficacy of targeted and immunotherapeutic treatment. Otherwise, no further surgical interventions will be performed. At one month following surgery, patients will be administered camrelizumab (200 mg/dose, i.v. drip, q3w) combined with apatinib (250 mg, p.o., qd) if their liver function has returned to Child-Pugh grade A. If not, medication will be postponed for a maximum of 2 months to await the return of normal liver function. If liver function fails to reach normal levels within this interval, patients will be discharged from the trial.

> Patients will undergo treatment until day 90 following the first use of camrelizumab, or until disease progression or unacceptable toxicity manifest. When unacceptable toxicities are deemed to be apatinib-related by study investigators, the dosage can be reduced or discontinued as appropriate. However, no change in the camrelizumab dose will be permitted. If patients exhibit a grade ≥ 3 AE that is deemed by the investigator to be camrelizumab-related, this drug can be temporarily discontinued until the AE grade is ≤ 1. Camrelizumab may be suspended for a maximum of 3 weeks, and treatment can resume if symptoms recover during this interval.
>
> Otherwise, camrelizumab use will be permanently discontinued. This drug will also be permanently discontinued if patients exhibit AEs exceeding the following levels including but not limited to: grade 2 pneumonia, grade 2 or 3 diarrhea or enterocolitis, grade 3 dermatitis, grade 2 AST or ALT or TBIL with a duration < 7 days, grade 2 pituitary inflammation, grade 2 adrenocortical insufficiency, grade 2 hyperthyroidism, grade 3 hyperglycemia; grade 2 or grade 3 Cr elevation, grade 2 neurotoxicity, or other grade 3 AE first presentation.

### Evaluation and Follow-up

Patients will be screened to ensure that they meet eligibility criteria, after which informed consent will be taken. Demographic characteristics, past medical history, history of medication use, and physical examination will then be completed, including measurements of height, weight, temperature, respiration, blood pressure, and heart rate. ECOG PS scores, liver function, renal function, blood counts, and thyroid function will also be evaluated.

Participants will undergo follow-up for a minimum of 24 months following the end of treatment, with follow-up being performed every 2-3 months until endpoint events are reached or the trial is completed. Follow-up will include: routine blood tests, liver function testing, adenocarcinoma marker panels, hepatitis B virus (HBV) DNA quantification, ultrasound imaging of the liver/pancreas/bile ducts/spleen, contrast-enhanced computed tomography (CT) scans (cranial, thoracic, upper and lower abdomen), and additional contrast-enhanced magnetic resonance imaging (MRI) scans as appropriate. Bone emission computed tomography (ECT) will additionally be performed in patients harboring bone metastases.

### Study endpoints

Patient OS is the primary endpoint for this study, and it is defined by the interval between hepatectomy and all-cause death. Secondary endpoints include PFS, ORR, DCR, DOR, and TTR of extrahepatic target lesions.

### Data collection

The following data will be collected from patient medical records by a study assistant:

1. Gender and age.
2. Vital signs of parameters measured during routine physical examination, including height, weight, temperature, respiration, heart rate, and blood pressure.
3. Medical and family history.
4. Personal history of smoking, alcohol intake, and medication usage.
5. Laboratory test results including hematological indices, kidney function indices, liver function indices, thyroid function indices, and cancer-related indicators.
6. Results of imaging tests including liver MRI/CT scans, chest CT scans, isotope bone scans, liver ultrasound, and ECG results.

Schedule of assessments is detailed in table 1.

### Statistical methods

#### Sample size estimation

The 2-year OS rate for advanced HCC patients with Child-Pugh grade A liver function in the RESCUE trial that underwent first-line treatment with camrelizumab and apatinib was 43.3%.^[28]^ Given that the progression of intrahepatic lesions is the primary cause of mortality in individuals with advanced HCC, hepatectomy-mediated removal of intrahepatic lesions is expected to improve OS by a minimum of 20% to 63.3% at 2 years. Given the established 24-month enrollment and 24-month follow-up periods, the false-positive and false-negative error values for this trial were respectively set to 0.05 and 0.1, with 15% lost-to-follow-up resulting in a calculated sample size of 62 patients.

### Outcome measure analyses

A statistical analysis plan (SAP) will be prepared based on the planned treatment scheme to analyze the resultant data, with the primary analytical methods being described below.

### Endpoint measurement analysis

The main study goal is to determine if a combination of targeted and ICI-based treatment can extend the survival of advanced HCC patients following the removal of the bulk of the immune-resistant intrahepatic tumor tissue. OS and PFS will be analyzed with Kaplan-Meier curves, hazard ratios (HRs), and 95% confidence intervals (CIs). Multivariate regression analyses will be used to identify factors that may influence patient prognosis based on those factors significant (P < 0.05) in univariate analyses. All analyses will initially be performed on an intention-to-treat (ITT) bases, followed by sensitivity analyses performed on a per-protocol basis. DCR, DOR, ORR, and TTR will be compared with Fisher’s exact test. Safety outcomes will also be assessed. Parametric or nonparametric tests will be used for quantitative data, while Fisher’s exact probability test will be used for categorical data. A two-sided P < 0.05 will be considered significant.

### Interim analysis

An interim study analysis will be performed every three months following the beginning of recruitment, with study-related data being reported to an independent data monitoring committee (DMC). Reported data will include information pertaining to recruitment, case report form (CRF) recovery rates, data quality, protocol deviations, patient dropout rates, patient characteristics, treatment, toxicity events, and primary/secondary endpoint measures.

### Terminal analysis

The trial will be completed within 24 months of patient recruitment, with final analyses being conducted after all patients have initiated treatment and undergone follow-up for a minimum of 24 months.

### Patient and public involvement

Our study’s design, conduct, reporting, or distribution strategies were not influenced by patients or the general public.

### Adverse events and scheme adjustment

#### Adverse events

For this trial, AEs will include any of the following:

1. Any suspected adverse drug reactions.
2. All reactions due to drug overdose, abuse, allergy, or toxicity.
3. Any apparently unrelated illnesses, including the exacerbation of preexisting conditions.
4. Injuries or accidents, with a note being made regarding the resultant outcomes.
5. Any abnormalities detected upon physical/physiological examination that require further investigation or clinical treatment.
6. Any hepatectomy-, apatinib-, or camrelizumab-related complications. All adverse medical events that occur between the time at which subjects sign the informed consent form and the last study visit will be classified as AEs, irrespective of whether these events are related to study treatment. When AEs occur, reasons for treatment cessation must be noted in detail in that patient’s CRF. All patients, including those exhibiting poor compliance, should continue to follow the study protocol unless they exit the trial. Following interruptions in trial therapies, investigators will determine an appropriate follow-up plan based on that patient’s current circumstances. Participants can exit the trial at any time without penalty, or can withdraw following investigator re-evaluation.

## Ethics and dissemination

### Research ethics approval

The study protocol, informed consent form and other submitted documents were reviewed and approved by the Ethics Committee of the Guangxi Medical University’s Affiliated Cancer Hospital and for Human Study (reference number KS2022[124]).

### Trial exit

All participants will be allowed to withdraw from the trial at any time without restrictions, or may be asked to withdraw as investigators deem appropriate/necessary. The following reasons are causes for study withdrawal from the investigator’s perspective:

1. Unacceptable treatment-associated toxicity.
2. Any unforseen events for which investigators do not recommend further treatment.
3. Serious violations of the study protocol, including non-compliance or repeated absences.
4. Elimination by the investigator for clinical reasons unrelated to study treatment.

### Confidentiality

All patient documentation will be recorded and preserved in strict confidence in accordance with the Protection of Personal Data Act’s strong data security requirements (draft). Investigators are required to maintain secrecy for all materials not presented to the trial office, including screening lists. Complete trial records may be made available to regulatory inquiries or in extraordinary circumstances, provided the privacy of patients is protected. The Trial Office will ensure that all patient information remains strictly confidential, with no identifiable information being released to any third parties. The representatives of the Trial Office may access patient information as necessary for quality control objectives, provided patient privacy is protected.

### Dissemination policy

After the trial or analysis, the results will be submitted to a peer-reviewed publication, and the paper will be compiled by the Trial Management Group (TMG), with copyright distribution chosen jointly by the TMG.

## Data Availability

All data produced in the present study are available upon reasonable request to the authors

## Patient consent for publication

Obtained.

## REFERENCES

[1] Sung H, Ferlay J, Siegel RL, et al. Global Cancer Statistics 2020: GLOBOCAN Estimates of Incidence and Mortality Worldwide for 36 Cancers in 185 Countries[J]. CA Cancer J Clin, 2021, 71(3): 209–249.

[2] Zhong JH, Peng NF, You XM, et al. Tumor stage and primary treatment of hepatocellular carcinoma at a large tertiary hospital in China: A real-world study[J]. Oncotarget, 2017, 8(11): 18296–18302.

[3] Yau T, Tang VY, Yao TJ, et al. Development of Hong Kong Liver Cancer staging system with treatment stratification for patients with hepatocellular carcinoma[J]. Gastroenterology, 2014, 146(7): 1691-1700.e1693.

[4] Farinati F, Vitale A, Spolverato G, et al. Development and Validation of a New Prognostic System for Patients with Hepatocellular Carcinoma[J]. PLoS Med, 2016, 13(4): e1002006.

[5] Chan SL, Chong CC, Chan AW, et al. Management of hepatocellular carcinoma with portal vein tumor thrombosis: Review and update at 2016[J]. World J Gastroenterol, 2016, 22(32): 7289–7300.

[6] Xiang X, Lau WY, Wu ZY, et al. Transarterial chemoembolization versus best supportive care for patients with hepatocellular carcinoma with portal vein tumor thrombus:a multicenter study[J]. Eur J Surg Oncol, 2019, 45(8): 1460–1467.

[7] Zhou J, Sun H, Wang Z, et al. Guidelines for the Diagnosis and Treatment of Hepatocellular Carcinoma (2019 Edition)[J]. Liver Cancer, 2020, 9(6): 682–720.

[8] EASL Clinical Practice Guidelines: Management of hepatocellular carcinoma[J]. J Hepatol, 2018, 69(1): 182–236.

[9] Heimbach JK, Kulik LM, Finn RS, et al. AASLD guidelines for the treatment of hepatocellular carcinoma[J]. Hepatology, 2018, 67(1): 358–380.

[10] Vogel A, Cervantes A, Chau I, et al. Hepatocellular carcinoma: ESMO Clinical Practice Guidelines for diagnosis, treatment and follow-up[J]. Ann Oncol, 2018, 29(Suppl 4): iv238–iv255.

[11] Omata M, Cheng AL, Kokudo N, et al. Asia-Pacific clinical practice guidelines on the management of hepatocellular carcinoma: a 2017 update[J]. Hepatol Int, 2017, 11(4): 317–370.

[12] 2018 Korean Liver Cancer Association-National Cancer Center Korea Practice Guidelines for the Management of Hepatocellular Carcinoma[J]. Gut Liver, 2019, 13(3): 227–299.

[13] Shao YY, Wang SY and Lin SM. Management consensus guideline for hepatocellular carcinoma: 2020 update on surveillance, diagnosis, and systemic treatment by the Taiwan Liver Cancer Association and the Gastroenterological Society of Taiwan[J]. J Formos Med Assoc, 2021, 120(4): 1051–1060.

[14] Llovet JM, Ricci S, Mazzaferro V, et al. Sorafenib in advanced hepatocellular carcinoma[J]. N Engl J Med, 2008, 359(4): 378–390.

[15] Cheng AL, Kang YK, Chen Z, et al. Efficacy and safety of sorafenib in patients in the Asia-Pacific region with advanced hepatocellular carcinoma: a phase III randomised, double-blind, placebo-controlled trial[J]. Lancet Oncol, 2009, 10(1): 25–34.

[16] Kudo M, Finn RS, Qin S, et al. Lenvatinib versus sorafenib in first-line treatment of patients with unresectable hepatocellular carcinoma: a randomised phase 3 non-inferiority trial[J]. Lancet, 2018, 391(10126): 1163–1173.

[17] Casadei-Gardini A, Scartozzi M, Tada T, et al. Lenvatinib versus sorafenib in first-line treatment of unresectable hepatocellular carcinoma: An inverse probability of treatment weighting analysis[J]. Liver Int, 2021, 41(6): 1389–1397.

[18] Facciorusso A, Tartaglia N, Villani R, et al. Lenvatinib versus sorafenib as first-line therapy of advanced hepatocellular carcinoma: a systematic review and meta-analysis[J]. Am J Transl Res, 2021, 13(4): 2379–2387.

[19] Qin S, Bi F, Gu S, et al. Donafenib Versus Sorafenib in First-Line Treatment of Unresectable or Metastatic Hepatocellular Carcinoma: A Randomized, Open-Label, Parallel-Controlled Phase II-III Trial[J]. J Clin Oncol, 2021, 39(27): 3002–3011.

[20] El-Khoueiry AB, Sangro B, Yau T, et al. Nivolumab in patients with advanced hepatocellular carcinoma (CheckMate 040): an open-label, non-comparative, phase 1/2 dose escalation and expansion trial[J]. Lancet, 2017, 389(10088): 2492–2502.

[21] Finn RS, Ikeda M, Zhu AX, et al. Phase Ib Study of Lenvatinib Plus Pembrolizumab in Patients With Unresectable Hepatocellular Carcinoma[J]. J Clin Oncol, 2020, 38(26): 2960–2970.

[22] Finn RS, Qin S, Ikeda M, et al. Atezolizumab plus Bevacizumab in Unresectable Hepatocellular Carcinoma[J]. N Engl J Med, 2020, 382(20): 1894–1905.

[23] Lee MS, Ryoo BY, Hsu CH, et al. Atezolizumab with or without bevacizumab in unresectable hepatocellular carcinoma (GO30140): an open-label, multicentre, phase 1b study[J]. Lancet Oncol, 2020, 21(6): 808–820.

[24] Ren Z, Xu J, Bai Y, et al. Sintilimab plus a bevacizumab biosimilar (IBI305) versus sorafenib in unresectable hepatocellular carcinoma (ORIENT-32): a randomised, open-label, phase 2-3 study[J]. Lancet Oncol, 2021, 22(7): 977–990.

[25] Chen K, Wei W, Liu L, et al. Lenvatinib with or without immune checkpoint inhibitors for patients with unresectable hepatocellular carcinoma in real-world clinical practice[J]. Cancer Immunol Immunother, 2022, 71(5): 1063–1074.

[26] Deng ZJ, Li L, Teng YX, et al. Treatments of Hepatocellular Carcinoma with Portal Vein Tumor Thrombus: Current Status and Controversy[J]. J Clin Transl Hepatol, 2022, 10(1): 147–158.

[27] Qin S, Li Q, Gu S, et al. Apatinib as second-line or later therapy in patients with advanced hepatocellular carcinoma (AHELP): a multicentre, double-blind, randomised, placebo-controlled, phase 3 trial[J]. Lancet Gastroenterol Hepatol, 2021, 6(7): 559–568.

[28] Xu J, Shen J, Gu S, et al. Camrelizumab in Combination with Apatinib in Patients with Advanced Hepatocellular Carcinoma (RESCUE): A Nonrandomized, Open-label, Phase II Trial[J]. Clin Cancer Res, 2021, 27(4): 1003–1011.

[29] Lam CM, Lo CM, Yuen WK, et al. Prolonged survival in selected patients following surgical resection for pulmonary metastasis from hepatocellular carcinoma[J]. Br J Surg, 1998, 85(9): 1198–1200.

[30] Chan KM, Yu MC, Wu TJ, et al. Efficacy of surgical resection in management of isolated extrahepatic metastases of hepatocellular carcinoma[J]. World J Gastroenterol, 2009, 15(43): 5481–5488.

[31] Yang T, Lu JH, Lin C, et al. Concomitant lung metastasis in patients with advanced hepatocellular carcinoma[J]. World J Gastroenterol, 2012, 18(20): 2533–2539.

[32] Kow AW, Kwon CH, Song S, et al. Clinicopathological factors and long-term outcome comparing between lung and peritoneal metastasectomy after hepatectomy for hepatocellular carcinoma in a tertiary institution[J]. Surgery, 2015, 157(4): 645–653.

[33] Zhong JH, Ke Y, Gong WF, et al. Hepatic resection associated with good survival for selected patients with intermediate and advanced-stage hepatocellular carcinoma[J]. Ann Surg, 2014, 260(2): 329–340.

[34] Yuan BH, Yuan WP, Li RH, et al. Propensity score-based comparison of hepatic resection and transarterial chemoembolization for patients with advanced hepatocellular carcinoma[J]. Tumour Biol, 2016, 37(2): 2435–2441.

[35] Llovet JM, De Baere T, Kulik L, et al. Locoregional therapies in the era of molecular and immune treatments for hepatocellular carcinoma[J]. Nat Rev Gastroenterol Hepatol, 2021, 18(5): 293–313.

[36] Galluzzi L, Buqué A, Kepp O, et al. Immunogenic cell death in cancer and infectious disease[J]. Nat Rev Immunol, 2017, 17(2): 97–111.

[37] Cheng AL, Hsu C, Chan SL, et al. Challenges of combination therapy with immune checkpoint inhibitors for hepatocellular carcinoma[J]. J Hepatol, 2020, 72(2): 307–319.

[38] Ngwa W, Irabor OC, Schoenfeld JD, et al. Using immunotherapy to boost the abscopal effect[J]. Nat Rev Cancer, 2018, 18(5): 313–322.

[39] Reig M, Forner A, Rimola J, et al. BCLC strategy for prognosis prediction and treatment recommendation: The 2022 update[J]. J Hepatol, 2022, 76(3): 681–693.

