## Supplementary material for "Hepatectomy combined with targeted and immunotherapy for CNLC stage IIIb hepatocellular carcinoma: a single-arm clinical trials protocol": Table 1

| Table 1 Assessments and time points | | | | |
| --- | --- | --- | --- | --- |
| Assessment | Time points | | | |
|  | ≤7 days before enrollment | beginning of PD-1 cycle 2 (day 22) | beginning of PD-1 cycle 5 (day 85) | Days of Cycle |
| routine blood test | X | X | X | X |
| routine urine test | X | X | X | X |
| liver function test | X | X | X | X |
| kidney function test | X | X | X | X |
| thyroid function test | X |  | X | X |
| myocardial enzyme | X |  | X | X |
| cellular immunity | X | X | X | X |
| HBV DNA | X |  | X | X |
| coagulation function | X | X | X | X |
| AFP | X | X | X | X |
| CA199 | X |  | X | X |
| ECG | X |  | X | X |
| Colour Doppler ultrasound | X |  | X | X |
| contrast-enhanced liver MRI/CT | X |  | X | X |
| thoracic CT | X |  | X | X |
| radionuclide bone scan | X |  |  | X |
| Additional imaging | X |  | X | X |

ECG, electrocardiogram;

Days of Cycle, every 3 cycles after the 5th cycle until tumor progression or intolerance to treatment;

Additional imaging, imaging examination of the other metastatic;
